# Medical tourism’s vulnerability to COVID-19 and its path to recovery: a 10-year data analysis of international patient visits

**DOI:** 10.1101/2023.10.26.23297607

**Authors:** Gihyun Myung, Juwon Lim

## Abstract

**Background:** This study seeks to assess the trends in international patient intake over the past decade, with a specific focus on the impact of the COVID-19 pandemic.

**Methods:** This is a retrospective study on the data obtained from SUPREME (CDW Research Search System) of Information Center from a single university hospital. Patient demographics on the gender, age, nationality, and diagnosis visiting International Healthcare Center from January 2013 to June 2023 were collected, and the impact and recovery rates from COVID-19 were calculated in terms of the number of foreign patients. The patient number was further analyzed based on gender, age, nationality, and diagnosis.

**Results:** The impact and recovery rates of COVID-19 on the number of foreign patients was 57% and 71%, respectively. The patients from USA, the old adult group (age 40-64), and patients with cancer were least affected by COVID-19, whereas the patients from the UAE, the young adult group (age 19-39), and patients for medical screening were most affected by COVID-19.

**Conclusions:** The number of international patients steadily was steadily rising since 2013 with a drop in number more than half with the advent of COVID-19 in 2019. However, recent data suggests a revival of this trend, signaling a gradual return to pre-pandemic levels. Despite the overall recovery, future trends are not entirely predictable due to potential factors. Future pandemics, international conflicts, or economic instability could potentially affect the influx of foreign patients. To revive medical tourism, a joint effort from the government and hospital is necessary.

## I. INTRODUCTION

International travel to seek medical care has many benefits including access to advanced technology, cost-saving, and reduced wait time.^1-3^ The number of international patients had been steadily growing in South Korea. In 2009, around 60,000 patients visited Korea for medical care. In 2017, the number increased to 320,000.^4^ The outbreak of COVID-19 in 2019 challenged healthcare workers, patients, and healthcare system worldwide. Coping with these challenges helped reshape the healthcare system. Most notably, telemedicine services have emerged to reduce the increased burden from COVID-19 and to prevent the spread of COVID-19.^5^ Efforts from the government and hospitals have successfully controlled the disease, and now even though not eradicated, we are entering the post-COVID era.

The COVID-19 pandemic has had a profound effect on international travel in Korea. It is not new though, and the sharp decrease was seen previously from SARS and MERS drastically reducing the number of international travels to South Korea in 2002 and 2015.^3^ In 2015, the total number of tourists travelling to Korea was 13,231,651, and the trend showed a sharp decrease from June to September and recovery to the previous level after October.^6^ As the pandemic ebbs, there are indicators pointing to a revival of medical tourism. For example, the number of foreign patients travelling to Korea dropped from 497,000 to 117,000 in 2020, but the number has increased to 145,000 in 2021 and 248,000 in 2022.^7^ Despite these positive signs, considerable uncertainties persist for the future, necessitating consistent monitoring and adaptability.

## II. METHODS

We utilized the SUPREME (CDW Research Search System) of the Seoul National University Hospital (SNUH) Information Center to collect data on foreign patients who visited the International Healthcare Center from January 2013 to June 2023. The data included patient demographics such as patient gender, age, nationality, and diagnosis, totaling 45,929 patient visits.

The impact of COVID-19 on the number of foreign patients was calculated by dividing the decrease in the number of patients in the post-COVID-19 year (2020) by the number of patients in the pre-COVID-19 year (2019). The recovery rate was calculated by dividing the average number of patients in the COVID-19 recovery year (2022 and 2023) by the number of patients in the pre-COVID-19 year (2019). Higher impact rate indicates steeper decline in patient numbers. We categorized and compared the trends of international patient visiting Seoul National University Hospital by country, age, and diagnostic categories to identify various factors associated with impact and recovery rates. We conducted our statistical analysis using STATA 18.0 (Stata Corp., College Station, TX, USA) to ensure the accuracy of our findings. Ethics approval was waived because this is based on a retrospective study of archived information that is fully anonymized before being accessed. Thus, there was no informed written consent required for individuals whose information was collected. The anonymized data was accessed on July 25, 2023.

## RESULTS

A total of 45,929 international patients visited the SNUH between 2013 and 2023. Among them, 19,614 (42.7%) were men, and 26,315 (57.3%) were women (Table 1). The most common age group was 19 to 39 (38.5%), and the most represented country was China (27.1%), followed by the USA (18.1%), Mongolia (15.7%), Kazakhstan (5.2%), United Arab Emirates (5.9%), and Russia (3.8%) (Table 1).

**Table 1.** Baseline characteristics of overseas patients from 2013 to 2023.

|  | Number | % |
| --- | --- | --- |
| Gender |  |  |
| Men | 19614 | 42.7 |
| Women | 26315 | 57.3 |
| Age groups |  |  |
| 0-18 | 8595 | 18.7 |
| 19-39 | 17680 | 38.5 |
| 40-64 | 16059 | 35.0 |
| 65- | 3595 | 7.8 |
| Nationality |  |  |
| China | 12423 | 27.1 |
| USA | 8314 | 18.1 |
| Mongolia | 7220 | 15.7 |
| Kazakhstan | 2712 | 5.9 |
| UAE | 2386 | 5.2 |
| Russia | 1769 | 3.8 |
| Others | 11105 | 24.2 |
| Year |  |  |
| 2013 | 4273 | 9.3 |
| 2014 | 5464 | 11.9 |
| 2015 | 4717 | 10.3 |
| 2016 | 4671 | 10.2 |
| 2017 | 4653 | 10.1 |
| 2018 | 5390 | 11.7 |
| 2019 | 5867 | 12.8 |
| 2020 | 2503 | 5.5 |
| 2021 | 2485 | 5.4 |
| 2022 | 3465 | 7.5 |
| 2023 | 2441 | 5.3 |
| Total | 45929 | 100 |

SNUH, Seoul National University Hospital; UAE, United Arab Emirates

The number of international patients visiting SNUH decreased substantially from 2019 to 2020 (5,867 to 2,503). However, by 2022, the annual number of international patients recovered to 59% of the number in 2019. With gradual recovery, the number of patients is expected to reach 4,882 by the end of 2023 (Figure 1).

**Figure 1.**
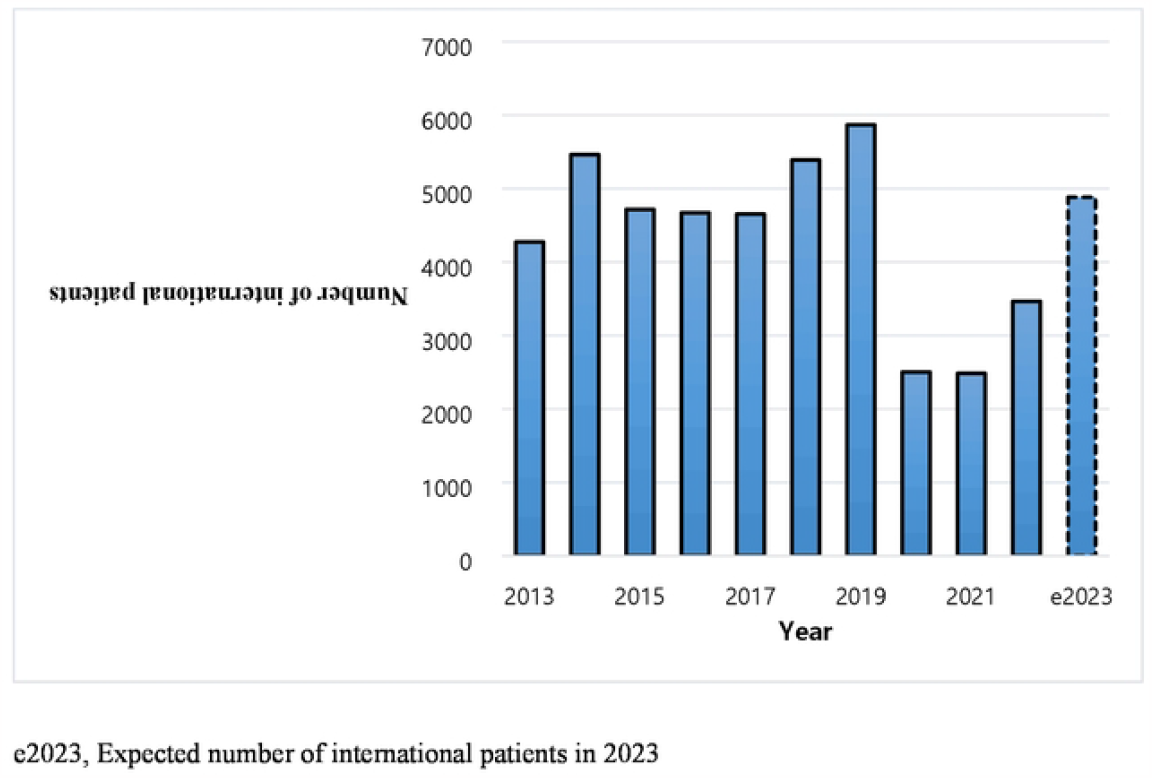
Trends in the number of international patients from 2013 to 2023. From 2013 to 2023, Seoul National University Hospital saw a sharp decline due to COVID-19, followed by a gradual recovery. By 2022, international patient visits recovered to 59% of the number in 2019. With gradual recovery, the number of patients is expected to reach 4,882 by the end of 2023. e2023, Expected number of international patients in 2023

The impact rate ranged from 33% to 77%; the USA was the least affected by the pandemic, whereas the UAE was the most impacted by the pandemic (Table 2). Generally, a low impact rate correlated with high recovery rate. The USA had the lowest impact rate (33%) followed by Russia (57%) and Mongolia (61%). The USA had the highest recovery rate (158%), which indicates that it is projected to exceed the pre-pandemic levels, followed by Mongolia (98%) and Russia (65%). However, countries like the UAE and China are only expected to reach between 38% and 31% of their previous patient numbers, respectively (Table 2).

**Table 2.** Impact and Recovery rate by nationality.

| Year | USA | China | Russia | Mongolia | UAE | Kazakhstan | Others | Total |
| --- | --- | --- | --- | --- | --- | --- | --- | --- |
| 2013 | 764 | 673 | 323 | 1138 | 280 | 198 | 897 | 4273 |
| 2014 | 868 | 1223 | 329 | 1092 | 383 | 419 | 1150 | 5464 |
| 2015 | 793 | 1185 | 192 | 792 | 292 | 445 | 1018 | 4717 |
| 2016 | 832 | 1340 | 153 | 629 | 281 | 350 | 1086 | 4671 |
| 2017 | 777 | 1253 | 171 | 661 | 320 | 301 | 1170 | 4653 |
| 2018 | 733 | 1857 | 159 | 749 | 301 | 352 | 1239 | 5390 |
| 2019 | 710 | 2403 | 171 | 708 | 264 | 329 | 1282 | 5867 |
| 2020 | 479 | 846 | 73 | 273 | 61 | 97 | 674 | 2503 |
| 2021 | 752 | 558 | 47 | 232 | 55 | 48 | 793 | 2485 |
| 2022 | 962 | 657 | 80 | 510 | 100 | 86 | 1070 | 3465 |
| e2023 | 1288 | 856 | 142 | 872 | 98 | 174 | 1452 | 4882 |
| Total | 8314 | 12423 | 1769 | 7220 | 2386 | 2712 | 11105 | 45929 |
| Impact rate | 33% | 65% | 57% | 61% | 77% | 71% | 47% | 57% |
| Recovery rate | 158% | 31% | 65% | 98% | 38% | 40% | 98% | 71% |

UAE, United Arab Emirates

Post-pandemic recovery rate differs by age. For example, the recovery rate for older patients, aged 65 and above, will significantly exceed the pre-pandemic level with an estimated increase of 123% compared to 2019. Similarly, the older adult group (age 40-64) is expected to recover to nearly 90% of their pre-pandemic level. However, younger adults, those between the ages of 19 and 39, are only anticipated to achieve a 50% recovery rate in comparison to their pre-pandemic numbers (Table 3).

**Table 3.**
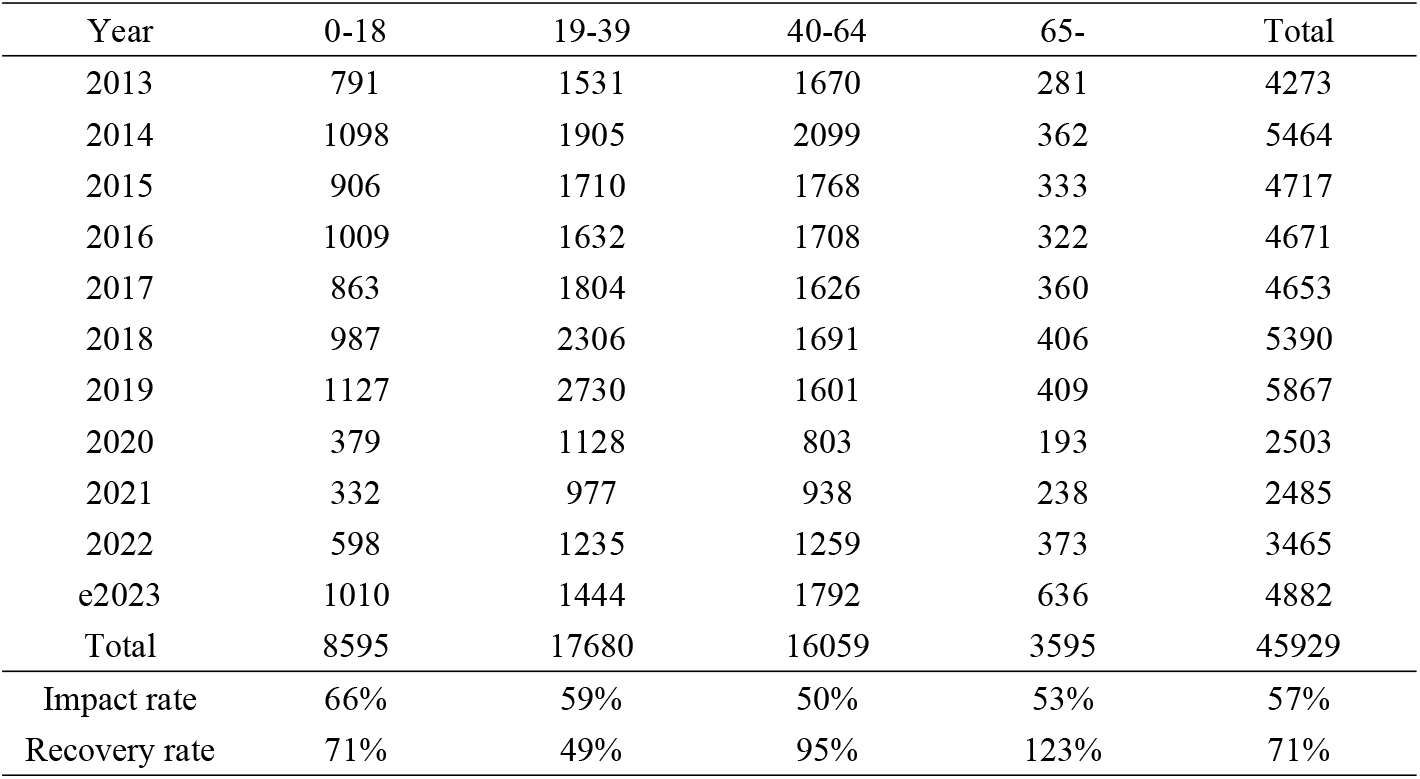
Impact and Recovery rates by age groups.

Based on disease categories, cancer, cardiovascular, and congenital conditions have recovery rates between 78% and 100% and projected to recover close to pre-pandemic levels (Table 4). On the contrary, patients visiting for a routine physical exam including for visas or diagnosed with infectious diseases are anticipated to achieve a recovery rate of less than half of their pre-pandemic levels, with the estimated recovery rate around 40%.

**Table 4.** Impact and Recovery rate by diagnostic categories.

| Year | Physical | Infection | Cancer | CVD | Congenital | Others | Total |
| --- | --- | --- | --- | --- | --- | --- | --- |
| 2013 | 549 | 386 | 403 | 202 | 102 | 2631 | 4273 |
| 2014 | 544 | 493 | 525 | 280 | 185 | 3437 | 5464 |
| 2015 | 576 | 390 | 427 | 197 | 142 | 2985 | 4717 |
| 2016 | 467 | 380 | 437 | 191 | 156 | 3040 | 4671 |
| 2017 | 571 | 339 | 237 | 107 | 85 | 3314 | 4653 |
| 2018 | 1211 | 334 | 201 | 133 | 84 | 3427 | 5390 |
| 2019 | 1730 | 371 | 197 | 146 | 98 | 3325 | 5867 |
| 2020 | 522 | 107 | 116 | 71 | 43 | 1644 | 2503 |
| 2021 | 655 | 45 | 102 | 44 | 37 | 1602 | 2485 |
| 2022 | 680 | 120 | 133 | 76 | 54 | 2402 | 3465 |
| e2023 | 684 | 220 | 262 | 178 | 98 | 3440 | 4882 |
| Total | 4430 | 2194 | 2731 | 1430 | 976 | 34168 | 45929 |
| Impact rate | 70% | 71% | 41% | 51% | 56% | 51% | 57% |
| Recovery rate | 39% | 46% | 100% | 87% | 78% | 88% | 71% |

## III. DISCUSSION

The COVID-19 pandemic has fundamentally reshaped the landscape of global medical tourism, with profound implications for healthcare providers and patients alike. Our study presents a comprehensive overview of the trends and projected recovery rates of international patient visits to SNUH. According to our data, the overall recovery rate is expected to reach 71% by the end of 2023. This indicates a promising, albeit slow, recovery from the COVID-19 pandemic. The signs of recovery are shown in foreign patients visiting Korea in general. According to the 2022 Foreign Patient Arrivals Statistical Analysis Report by the Korea Healthcare Industry Promotion Agency, there has been a noticeable recovery in the number of foreign patients visiting South Korea for healthcare needs following the slump induced by the COVID-19 pandemic.^8^ In 2022, the number of foreign patients visiting Korea totaled 24.8 million, indicating a significant rebound from the 11.7 million in 2020 and 14.5 million in 2021 during the peak of the pandemic. Breaking down the patient inflow by nationality, the majority came from the United States, followed by China, Japan, Thailand, and Vietnam in 2022.^9^

Our data indicates that the recovery rate differed by country, age, and diagnosis. These findings reveal that recovery process is complex and many different factors influence medical tourism.

First of all, countries have different travel restrictions and availability of healthcare services in their home countries. Countries like USA (including US Army from Pyeongtaek, South Korea) and Mongolia, which are projected to reach or exceed their pre-pandemic patient levels, may have favorable conditions promoting medical tourism. This aligns with the general trend where countries highly represented in medical tourism to Korea actually demonstrate an increased number of patients in 2022 compared to 2020 or 2021.^8^ This signals a general resurgence in global medical tourism.

In contrast, the slower recovery rates for China, Russia, and the UAE suggest ongoing challenges and barriers. Generally, shorter distance from Korea correlated with higher recovery rate except Russia and China (Figure 2). Inflation fueled by the war with Ukraine coupled with rising airfares and fewer flights pose challenges for international patients in Russia.^10^ Rising tensions between China and Korea have hindered the recovery of medical tourism from COVID-19 (Figure 2).^11^

**Figure 2.**
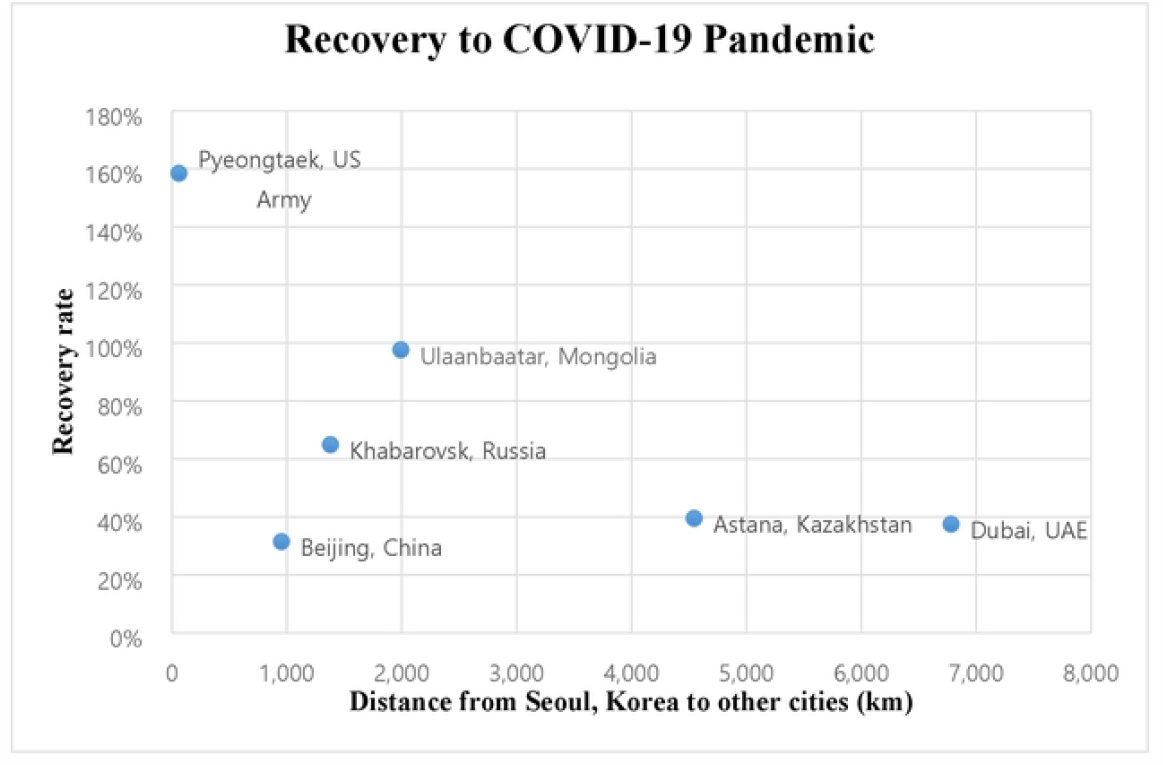
Resilience to COVID-19 Pandemic. Generally, shorter distance from Korea correlated with higher recovery rate except Russia and China.

To revive medical tourism, a collaborative effort between hospitals and government is necessary. The UAE now has emerged as a significant economic partner for South Korea, extending beyond traditional construction and energy industries to encompass private sectors as well. ^12^ Medical tourism can play a mediator role in expanding cooperation between two countries ranging from IT, renewable energy, and to defense.^12^

The two countries can utilize their common interest in health and medical cooperation to create new policies to facilitate easier and more efficient process for medical tourism. In addition, the government can help hospitals by offering incentives for medical tourism. As an example, Malaysian government offers tax incentives and expenses in receiving international accreditation such as Joint Commission international (JCI).^4,13^

Hospitals can utilize telehealth to facilitate smooth transition of care from the country of origin to South Korea by providing virtual care in cases such as pre-operative and post-operative care.

Interestingly, the recovery rates demonstrate differences in urgency and the availability of treatments for various conditions. The full recovery projected for diagnostic groups like cancer, cardiovascular disease, and congenital anomalies underscores the critical and urgent nature of these treatments. The lower recovery rates among patients seeking routine physical examinations or diagnosed with infectious diseases may be linked to the risk of cross-infection or the expanded capabilities of telehealth services during the pandemic.

Limitations of this study include that it is a single-center experience, which may not capture the full breadth of trends in medical tourism across South Korea. SNUH is one of the biggest academic centers in Korea with more than 800 hospital beds. Thus, we believe that our data can be a good representation of the overall trends. Another limitation is that we have data up to June 2023, and the expected number for 2023 is an estimation based on the 1^st^ half of 2023. Due to the possibility that the number from the second half of 2023 was undervalued, the actual recovery rate is expected to be higher.

## IV. CONCLUSION

Our study provides valuable insights into the recovery rates among foreign patients, further research is required to explore the underlying factors affecting these trends more deeply. Despite its limitations, we believe that our study makes a valuable contribution to understanding the impact of the COVID-19 pandemic on medical tourism and can serve as a foundation for future research and policy-making.

## Data Availability

All relevant data are within the manuscript and its Supporting Information files.

## Acknowledgements

There is no acknowledgement

## Author Contributions

GM and JL had contributions to the conception, data curation, methodology, data acquisition, formal analysis, interpretation of data, and provided draft and editing for the work, and final approval of the version

